# The motor anomalies seen in isolated REM sleep behavior disorder

**DOI:** 10.1101/2023.06.11.23291091

**Authors:** Cristina Simonet, Laura Pérez-Carbonell, Brook FR Huxford, Harneek Chohan, Aneet Gill, Guy Leschziner, Andrew J Lees, Anette Schrag, Alastair J Noyce

## Abstract

**Background:** Isolated REM sleep behavior disorder (iRBD) is known to be an early feature in some PD patients. Quantitative tools are needed to detect early motor anomalies in iRBD.

**Methods:** A motor battery was used to compare iRBD patients with controls. This included two online keyboard-based tests, the BRadykineisa Akinesia INcoordination (BRAIN) test and the Distal Finger Tapping (DFT) test, a timed handwriting task and two motor assessments (10-meter walking and finger tapping) carried out both alone and during a mental task. This battery was compared with the motor section of the MDS-MDS-UPDRS-III. ROC analyses were used to measure diagnostic accuracy.

**Results:** We included 33 patients with video-PSG-confirmed iRBD and 29 age and sex matched controls. The iRBD group performed the BRAIN test and DFT test more slowly (p<0.001, p=0.020 respectively) and erratically (p<0.001, p=0.009 respectively) than controls. Handwriting speed was 10 seconds slower in iRBDs than controls (p=0.004). Unlike controls, under a mental task the iRBD group decreased their walking pace (p<0.001) and had a smaller amplitude (p=0.001) and slower (p=0.007) finger tapping than tasks in isolation. The combination of BRAIN & DFT tests with the effect of mental tasks on walking and finger tapping showed 90.3% sensitivity for 89.3% specificity (AUC 0.94, 95% CI 0.88-0.99), which was higher than the MDS-UPDRS-III (minus action tremor) (69.7% sensitivity, 72.4% specificity; AUC 0.81, 95% CI 0.71-0.91) for detecting motor abnormalities.

**Conclusion:** This study suggests that speed, incoordination, and dual task motor deterioration might be accurate indicators of incipient PD in iRBD.

## Introduction

Isolated rapid eye movement (REM) sleep behavior disorder (iRBD) is a parasomnia characterized by the presence of vivid dreams and acting-out behaviors that occur during REM sleep, with demonstrated loss of REM atonia by video-polysomnography (v-PSG).^1^ In patients with iRBD, pathological α-synuclein may be found in the cerebrospinal fluid and other tissues.^2, 3^ Deposits of α-synuclein have also been found in post-mortem brains of subjects with iRBD.^4^ Several prospective longitudinal studies have shown that more than 80% of patients with iRBD develop an α-synucleinopathy after 10 years.^5–7^ Although PD has been described to be the commonest final diagnosis in patients with iRBD, 43.5% patients with iRBD eventually developed Dementia with Lewy bodies and 4.5% Multiple System Atrophy.^7^ Annual conversion rates from iRBD to an α-synucleinopathy range between 6.3%^7^ and 8% per year.^5^

Motor dysfunction has been found to be the strongest predictive marker of conversion of future parkinsonism or dementia in patients with iRBD.^7^ Clinical rating scales are not designed for use in the early prodromal stages of PD and may not be sensitive enough to pick up subtle motor anomalies.^8^ Their floor effect and insensitivity in the earliest stages of disease are the most important limiting factors.^9^ A study of v-PSG-confirmed iRBD, carried out by the International RBD Study Group, followed up 1280 patients with iRBD over a median of 3.6 years and found that, unlike clinical rating scales, the combination of quantitative motor tests (alternate finger tapping, Timed up and Go test and Purdue Peg Board test) were the most powerful predictive marker of future diagnosis of an α-synucleinopathy (hazard ratio 3.16; 95% CI, 1.86 to 5.37)^7^.

Defining the natural progression of clinical markers during the prodromal stages of disease is important during the design of randomized trials in order to establish optimal clinical end points. The aim of this study was to develop an accurate and replicable battery of motor tests capable of identifying the most salient motor signatures in people with iRBD, and to determine the most sensitive motor markers of incipient parkinsonism to be followed over time.

## Material and methods

Patients with iRBD were identified from the Sleep Clinic at Guy’s Hospital and enrolled in the PREDICT-PD study.^10^ All patients had an overnight v-PSG that confirmed the diagnosis of iRBD. Healthy controls were recruited from lower risk PREDICT-PD study participants and were matched for age and sex with the iRBD group.

Exclusion criteria for iRBD and controls were having a formal diagnosis of dementia, PD, or other neurological conditions that could affect their motor performance such as essential tremor, motor neuron disease, multiple sclerosis, or polyneuropathy. We also checked for current medication with potential parkinsonian side effects.

We recruited 34 people with v-PSG-confirmed iRBD and 35 age- and sex-matched controls. Groups were also comparable with respect to medical comorbidities (Table 1). One patient with iRBD was not included in the final analysis due to fulfilling the diagnostic criteria for PD when seen in person. We excluded three controls with a diagnosis of essential tremor, two with rest tremor and rigidity on examination and one with cognitive impairment. In the end, we included 33 patients with iRBD and 29 age- and sex-matched controls in the analysis.

**Table 1.**
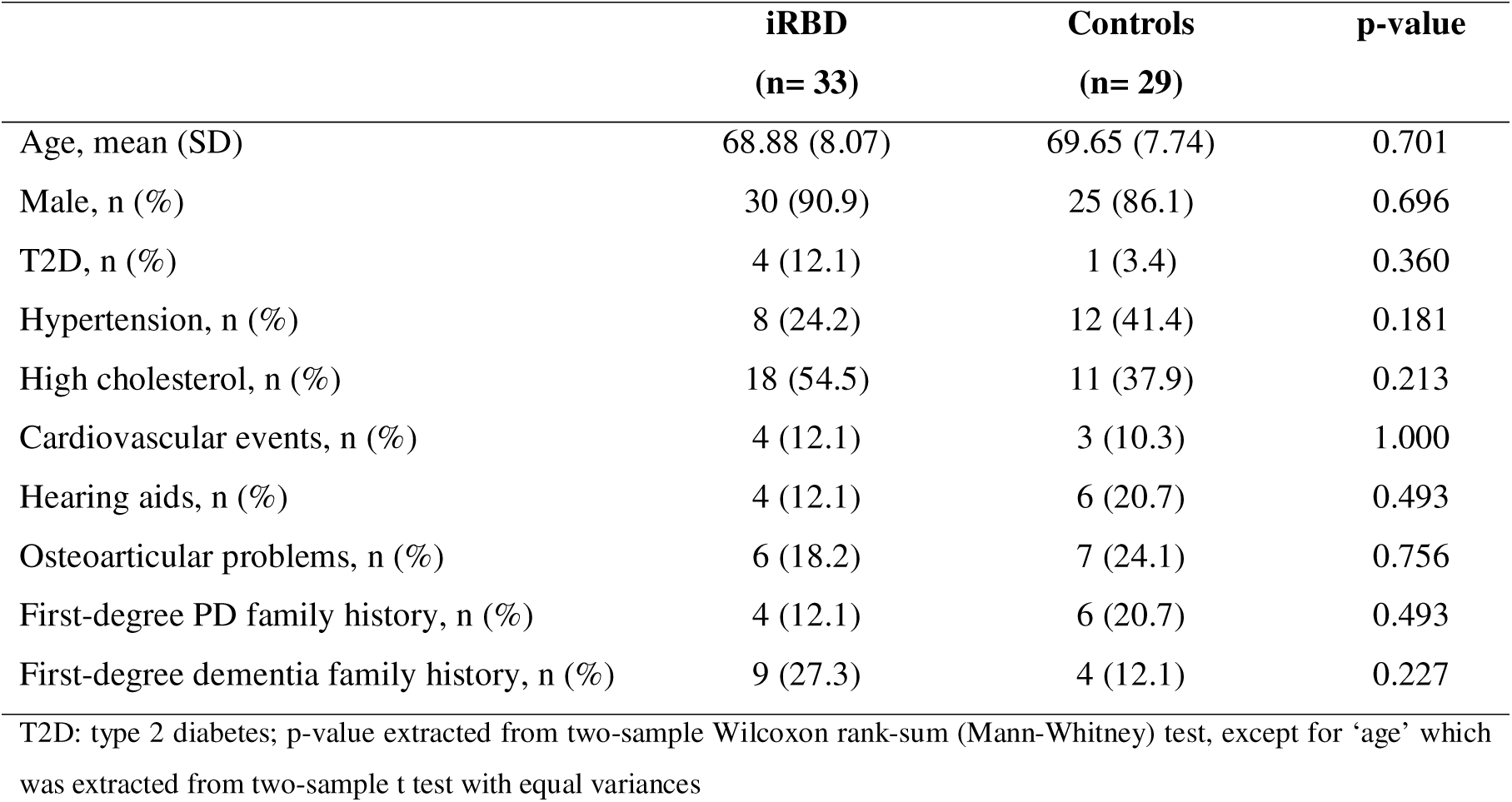
Demographic information, risk factors and non-motor manifestations.

|  | **iRBD**<br>**(n= 33)** | **Controls**<br>**(n= 29)** | **p-value** |
| --- | --- | --- | --- |
| Age, mean (SD) | 68.88 (8.07) | 69.65 (7.74) | 0.701 |
| Male, n (%) | 30 (90.9) | 25 (86.1) | 0.696 |
| T2D, n (%) | 4 (12.1) | 1 (3.4) | 0.360 |
| Hypertension, n (%) | 8 (24.2) | 12 (41.4) | 0.181 |
| High cholesterol, n (%) | 18 (54.5) | 11 (37.9) | 0.213 |
| Cardiovascular events, n (%) | 4 (12.1) | 3 (10.3) | 1.000 |
| Hearing aids, n (%) | 4 (12.1) | 6 (20.7) | 0.493 |
| Osteoarticular problems, n (%) | 6 (18.2) | 7 (24.1) | 0.756 |
| First-degree PD family history, n (%) | 4 (12.1) | 6 (20.7) | 0.493 |
| First-degree dementia family history, n (%) | 9 (27.3) | 4 (12.1) | 0.227 |

T2D: type 2 diabetes; p-value extracted from two-sample Wilcoxon rank-sum (Mann-Whitney) test, except for 'age' which was extracted from two-sample t test with equal variances

Participants were examined following the motor part of Movement Disorder Society-Unified Parkinson’s Disease Rating Scale (MDS-UPDRS-III) instructions.^11^ The MDS-UPDRS-III is used to apply the clinical criteria of Subthreshold Parkinsonism established by the MDS Task Force for Research Criteria for Prodromal PD which is based on MDS-UPDRS-III score >6 points after excluding action tremor.^12^ Given the cross-sectional nature of this study, the prevalence of Subthreshold Parkinsonism was used to explore the accuracy of motor assessments to distinguish between iRBD people with and without Subthreshold Parkinsonism.

Two movement disorders experts (AJL and AJN) independently scored the video recordings of participants following the instructions of the MDS-UPDRS-III. They were blind to the case/control status and did not have any additional information about participants. Given that rigidity could not be ascertained from the videos, we used rigidity scores from the in-person assessment.

### Motor battery

Five separate motor tests were included in the motor battery: two keyboard tapping tests (BRAIN and DFT), a timed-handwriting, a 10-meter walking test and a finger tapping task. The last two assessments were performed in isolation and under a mental task (Figure 1).

**Figure 1.**
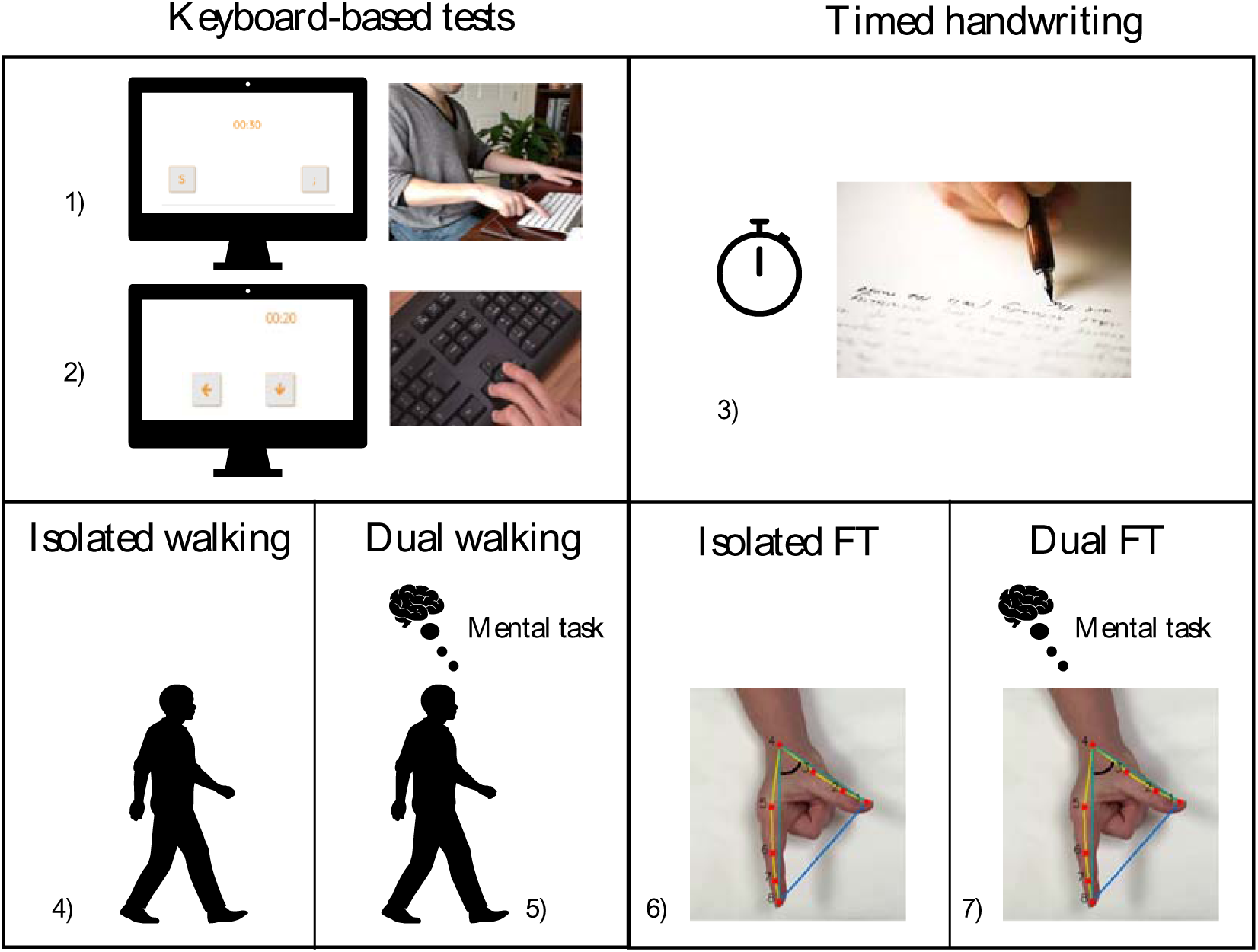
Motor battery description. 1) BRAIN (BRadykinesia Akinesia INcoordination) test, 2) DFT (Distal Finger Tapping), 3) timed handwriting task (writing *‘Mary had a little lamb, its fleece was white as snow’* three times using a pen and white paper), 4) 10-metre walking (timed), 5) 10-metre walking while doing a mental task (timed), 6) SMART test (Slow-Motion Analysis of Repetitive Tapping) in isolation and 7) SMART test in combination with a mental task

The BRadykinesia Akinesia INcoordination (BRAIN)^1^^3–15^ and Distal Finger Tapping (DFT)^16^ tests are web-based keyboard tapping tests which have been validated to remotely evaluate upper limb movements and quantify separate kinetic components such as speed, akinesia and rhythm as part of motor dysfunction in PD. They are available online and are compatible with regular laptops and computers with a keyboard. The BRAIN test is a 30- second alternating finger tapping test. The DFT test consists of a 20-second single key tapping test. Kinetic parameters of both tests include: 1) Kinesia Score (KS), which is a measure of speed (number of keystrokes typed in 30 seconds, (BRAIN test) or 20 seconds (DFT test)), 2) Akinesia Time (AT), which is a measure of akinesia (average dwell time (msec) that keys are depressed), and 3) Incoordination Score (IS), which measures the rhythm (variance (msec2) of travelling time between keystrokes). In addition, participants were timed while copying the sentence ‘*Mary had a little lamb, its fleece was white as snow*’ three times using a pen and white paper.

It is known that combining a cognitive activity with a motor task can unmask early motor dysfunction in individuals at risk of developing PD.^17^ We invited participants to perform two motor tasks (10-meter walking and finger tapping) in isolation and while doing a mental task (listing the months of the year in reverse order and subtracting ’3’ from ’100’ continuously, respectively). Finger tapping was assessed using the Slow-Motion Analysis of Repetitive Tapping (SMART) test which is a video-based tool focused on tracking repetitive finger tapping movements using a slow-motion camera. We invited patients to tap their index finger on the thumb as fast and as widely as they could following the same standardized instructions as the MDS-UPDRS-III (FT-sub-score). The finger tapping task was analyzed using a software to automatically detect the hand and process video images to analyze the finger tapping task.^18^ Two parameters were calculated: amplitude (angle between index finger and thumb) and velocity (distance travelled per second extracted from the derivative of the amplitude). For each parameter, we extracted the mean, SD and coefficient variation (CV) (SD/mean). In a previous study, the SMART test was shown to be accurate to distinguish patients with early PD (less than 2 years of disease duration) and also people with idiopathic anosmia who are known to be at risk to develop PD in the future from controls.^18^ For more detailed information about the test, see the supporting material.

### Statistical analysis

Data normality was assessed using the D’Agostino test. The mean and SD were calculated for normally distributed data. Median and interquartile range (IQR) were calculated for non-normal distributed data. Categorical variables were presented by absolute frequency and percentage and compared with Fisher’s exact test. Quantitative data for demographic and motor outcomes were compared using the Welch’s test for unequal variances.

For each motor test, we undertook one pairwise discriminatory comparison between participants with iRBD and controls. We used logistic regression and ROC curves to define AUC values for each quantitative motor marker and the MDS-UPDRS-III. We repeated the analysis using the combination of the most accurate motor parameters in the motor battery. Spearman’s rank correlation was used to determine whether motor parameters were independent or not. We included in the multivariate logistic model only those motor markers that were found to be independent to each other based on their correlation coefficient. The Wilson/Brown method was used to generate ROC curves and determine the accuracy (sensitivity and specificity of parameters) of each test in detecting iRBD patients from controls, iRBD with Subthreshold Parkinsonism from controls, and iRBD with and without Subthreshold Parkinsonism. A cut-off that maximized Youden’s J index was selected for each variable.

All statistical tests were two-tailed. We adjusted for multiple comparisons to avoid an increase in type 1 error. We applied the Bonferroni calculation to adjust the cut-off for evidence of association. The p-value was set at <0.010. Data analysis was carried out using STATA v.13 (StataCorp, College Station, TX).

Ethics approval was granted by the Queen Square Research Ethics Committee (09/H0716/48). Participants received verbal and written information about the study and appropriately consented to take part in the study.

## Results

Both groups were comparable in terms of age (mean (SD), iRBD: 68.88 years (8.07) vs controls: 69.65 years (7.74); p=0.701). Male predominance was present in the iRBD group (30/33 -90.9%-) and in the control group (25/29 -86.2%-; p=0.696). With respect to medical comorbidities, the iRBD group was comparable in terms of vascular risk factors including type 2 diabetes (T2D) (p=0.360), high cholesterol (p=0.213), hypertension (p=0.181), cardiovascular disease (p=1.000) and osteoarticular problems (p=0.756) (Table 1).

Patients with iRBD had a mean disease duration from symptom onset of 10.6 years (SD 6.87). Thirteen patients were taking symptomatic medication to treat iRBD (clonazepam and/or melatonin) and four were on treatment with selective serotonin reuptake inhibitors for depression. People with iRBD were more likely to have an abnormal cognitive test: eight iRBD participants scored less than 26 in the MoCA test (seven patients were classified as having mild cognitive impairment (25-22 points) and one as having moderate cognitive impairment (21-19 points)). In contrast, no controls had a score less than 26. Of note, both groups had similar years of education (iRBD 19.42 years (SD 6.35) vs controls 20.14 years (SD 3.31); p=0.575).

On average, motor scores differed by approximately 5 points (mean MDS-UPDRS-III (SD), iRBD: 7.24 (4.81); control: 2.65 (1.80); p<0.001). Blinded video analysis agreed (<5- point difference) with 90.6% (56/62) in-person examinations. Eleven people with iRBD fulfilled criteria for Subthreshold Parkinsonism following using the MDS Task Force criteria, which excludes action (intentional and postural) tremor. In contrast, none of the controls scored more than 6 points in the MDS-UPDRS-III, after excluding action tremor. One of the participants with Subthreshold Parkinsonism received the diagnosis of PD 8 months after our assessment (see the Box below for detailed clinical information).

### BOX: Newly diagnosed PD case

A male in his 60s with a more than 10-year history of iRBD. He did not have any motor symptoms when we saw him in person. However, he fulfilled criteria for Subthreshold Parkinsonism (total score in the MDS-UPDRS-III - after excluding action tremor - 13). On examination, he exhibited facial hypomimia and low volume speech. He also had mild bradykinesia in both hands although it was more noticeable in his left hand. He did not have rigidity. When walking, he exhibited reduced right arm swing. He did not have rest tremor but bilateral mild action tremor in both hands. Eight months after our assessment, he started noticing motor symptoms which included right hand tremor, walking difficulties and short-term memory symptoms. Looking back to his motor performance, his scores surpassed the cut-off established to distinguish iRBD with Subthreshold Parkinsonism in four tasks: KS-BRAIN 48 taps (cut-off ≤ 49 taps), IS-DFT 5346.8 sec2 (cut-off ≥ 2210 sec2), 10-metre dual walking task 15.7 sec (cut-off ≥ 8 sec). He scored 24/30 in the MoCA with deficits in the visuo-spatial and executive domains (2/5), and delayed recall (3/5).

Four participants had a suspected/possible early PD/parkinsonism based on our clinician global impression scale. Clinical impression was based on the presence of bradykinesia (mild decrement in amplitude/frequency in finger/toe tapping) and rigidity (mild rigidity). Only one participant had rest tremor. Neither of them had motor symptoms neither endorsed functional impairment.

The MDS Research Criteria for Prodromal PD showed a low sensitivity (42.4%) with high specificity (96.5%) to distinguish between iRBD and controls. Decreasing the cut-off down from 6 to 3 points improved the accuracy with 69.7% sensitivity with 72.4% specificity (AUC 0.81, 95% CI 0.71 to 0.91).

### Motor battery

The performance of dominant and non-dominant hand was comparable in both keyboard tests and finger tapping task. We focused our results on the non-dominant in both groups.

Firstly, the keyboard-based tapping tests (BRAIN and DFT) showed differences between iRBD and controls (Table 2). The iRBD group performed the alternate tapping task (BRAIN test) and single finger tapping task (DFT test) more slowly and erratically than controls. People with iRBD tapped on average 11.5 keys fewer than controls (mean KS- BRAIN (SD), 49.45 taps (15.19) vs 61.03 taps (9.98); p<0.001). Although KS-DFT was lower in the iRBD group, the difference between groups was less evident than in the BRAIN test (mean KS-DFT (SD), 83.26 taps (15.76) vs 90.58 taps (11.62); p=0.020). The iRBD group performed both tests more erratically than controls, based on a greater variance of travelling time between keystrokes (IS). In both tests, IS was significantly higher in the iRBD group (median IS-BRAIN (IQR), 5354.66 msec^2^ (2702.75 to 11478.53) vs 2375.19 msec^2^ (1640.95 to 3874.55); p<0.001; median IS-DFT (IQR), 2210.62 msec^2^ (1049.74 to 3265.58) vs 800.48 msec^2^ (329.57 to 1364.56); p=0.006). Although patients with iRBD spent slightly longer dwell time on each key (AT), the discrepancy between groups was not as noticeable as with other parameters (mean AT-BRAIN (SD), 131.43 msec (50.56) vs 109.86 msec (25.89); p=0.018; mean AT-DFT (SD), 110.70 msec (27.56) vs 102.64 msec (23.03); p=0.107).

**Table 2.**
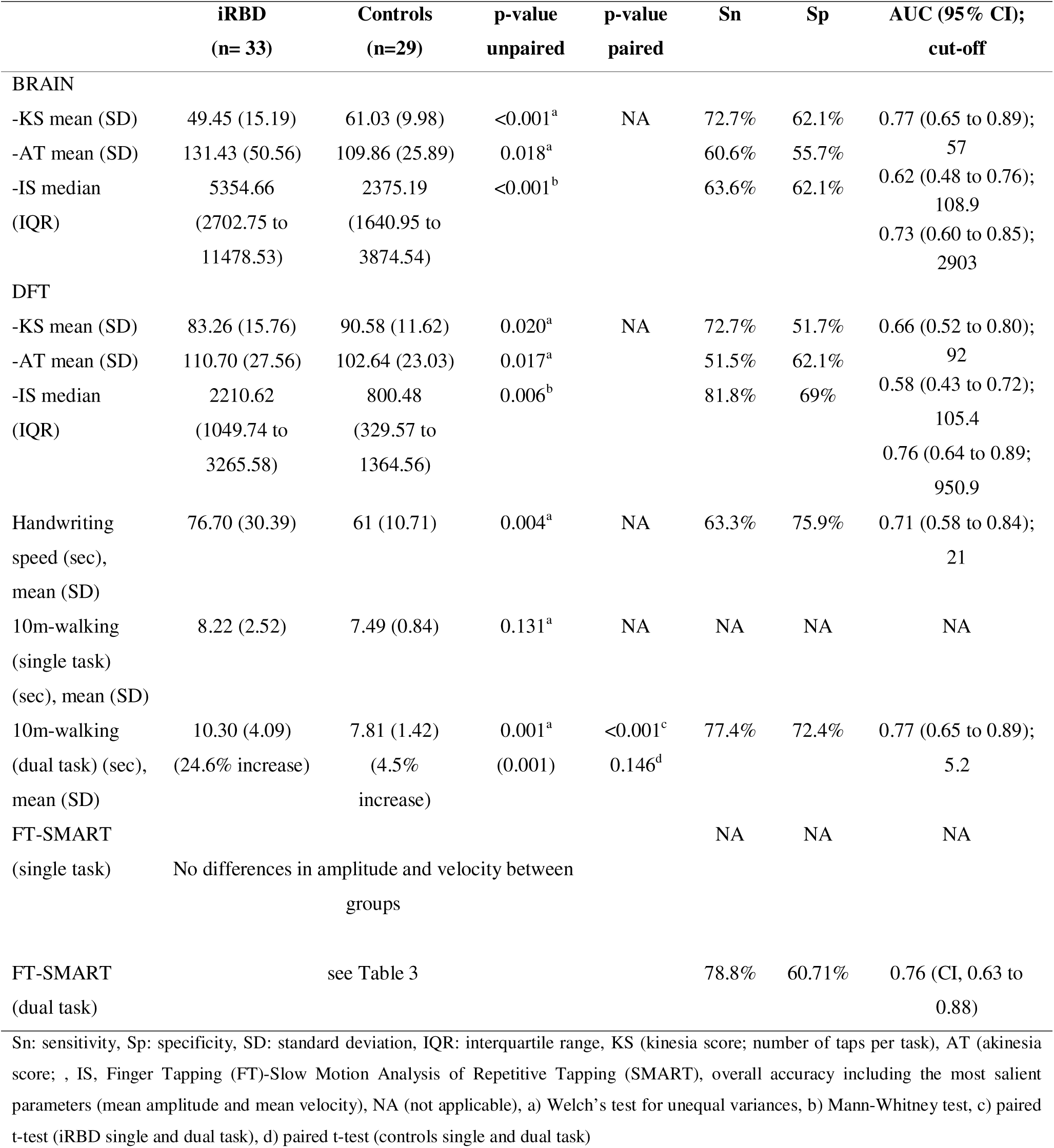
Motor battery: Group comparison and ROC analysis of salient motor markers.

All parameters discriminated between iRBD and controls. The number of alternate key taps (KS-BRAIN) and incoordination of single taps (IS-DFT) showed the best discriminatory power. The former had 72.7% sensitivity for 62.1% specificity (cut-off=57 taps) and AUC of 0.77 (95% CI 0.65 to 0.89). The latter had an 81.8% sensitivity for 69.0% specificity (cut-off=950.9 msec^2^) and AUC of 0.76 (95% CI 0.64 to 0.89). Both tests showed a high discriminatory power to detect iRBD with Subthreshold Parkinsonism amongst those without. The KS-BRAIN had 70% sensitivity for 73.9% specificity (cut-off=48 taps) and AUC of 0.72 (95% CI 0.49 to 0.96). The IS-DFT showed 80% sensitivity for 60.9% specificity (cut-off=2210 msec^2^) and AUC of 0.64 (95% CI 0.44 to 0.83) (Table 2).

Secondly, the iRBD group took 15 seconds longer to write three sentences than controls (mean time (SD), iRBD: 76.70 seconds (30.39) vs control 61 seconds (10.71); p=0.004). Handwriting speed was able to correctly detect iRBD patients from controls with 63.6% sensitivity for 75.9% specificity and using a cut-off of 21 seconds (AUC 0.71, 95% CI 0.58 to 0.84). When increasing the cut-off up to 24 seconds, it was found that handwriting speed was also able to accurately distinguish iRBD with Subthreshold Parkinsonism from those without Subthreshold Parkinsonism, with a detection rate of 81.8% for 82.8% specificity (AUC 0.86, 95% CI 0.68 to 1.00).

Thirdly, walking speed was similar (iRBD mean (SD): 8.22 seconds (2.52) vs controls mean (SD): 7.49 seconds (0.84); p=0.131) when carried out in isolation but differed amongst groups when it was performed under a mental task (iRBD mean (SD): 10.30 seconds (4.09) vs controls mean (SD): 7.81 seconds (1.42); p=0.001) (Table 2, Figure 2). Patients with iRBD slowed down their walking pace to a greater extent than controls (25% vs 4% increase of their walking time; p<0.001). The relative change between dual and single task duration (relative change (%): (dual task – single task)/single task * 100) was able to differentiate iRBD patients from controls with a 77.4% sensitivity and 72.4% specificity (AUC 0.77, 95% CI 0.65 to 0.89).

**Figure 2.**
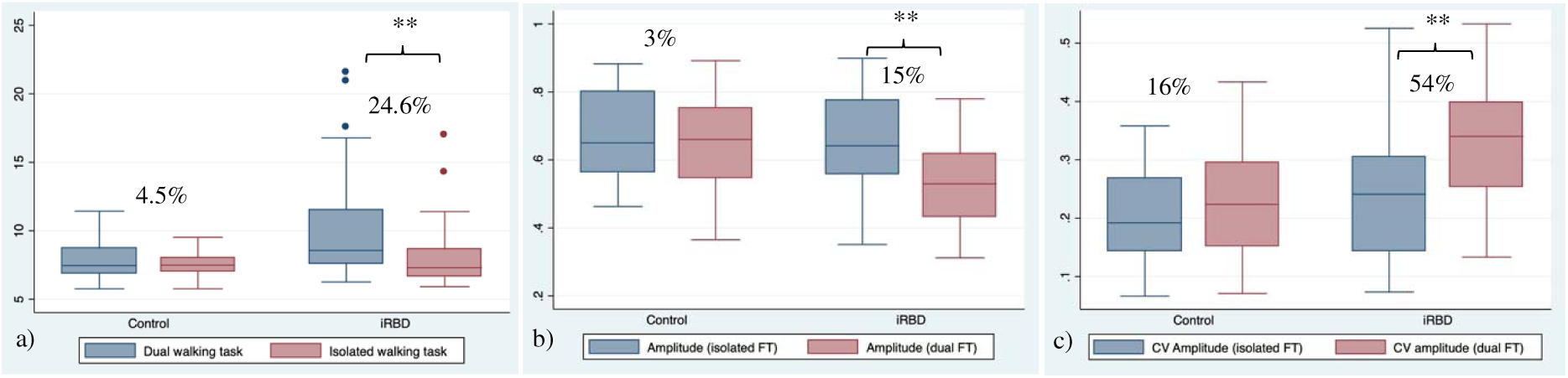
The effect of a mental task on walking speed (a), the mean amplitude of finger tapping (b), and the CV amplitude of finger tapping (c) ** p<0.001, % relative change (dual task – single task)/single task * 100. P-value calculated from a paired t-test

Finally, slow-motion analysis of repetitive finger tapping was similar in iRBD and in controls under natural conditions (isolated finger tapping). Unlike controls, dual tasking unmasked motor anomalies in people with iRBD. Under a mental task condition, the iRBD group performed the finger tapping task with a smaller amplitude (iRBD mean amplitude (SD): 0.54 (0.13) vs control mean amplitude (SD): 0.65 (0.14); p=0.001) and slower finger tapping (iRBD velocity mean (SD): 1.89×10^-2^ (5×10^-3^) vs control velocity mean: 2.23×10^-2^ (5×10^-3^); p=0.007) than controls (Table 3). People with iRBD decreased 15% their finger tapping amplitude when performing finger tapping under a cognitive task, whereas controls decreased only 3% (p=0.008). Finger tapping was noticeably more erratic (higher CV amplitude) in the iRBD group who increased 54% their CV compared with 16% increase in the control group (p<0.001) (Figure 2). There was a nominal evidence that iRBD patients slowed down their finger tapping velocity to a greater extent than controls (11% vs 9%; p=0.418). The CV of amplitude showed the highest accuracy to distinguish iRBD patients from controls (75.8% sensitivity for 64.3% specificity AUC 0.75, 95% CI 0.63 to 0.87) (Table 3).

**Table 3.** SMART test performance in isolation and under a mental task.

| **Single finger tapping** | **iRBD**<br>**(n=32) \*** | **Controls**<br>**(n=27) \*** | **p-value** |  | **Sn** | **Sp** | **AUC**<br>**(95% CI); cut-off** |
| --- | --- | --- | --- | --- | --- | --- | --- |
| Amplitude (a.u.) |  |  |  |  |  |  |  |
| -Mean | 0.65 | 0.68 | 0.541 |  | NA | NA | NA |
| -CV (SD/mean) | 0.23 | 0.20 | 0.189 |  |  |  |  |
| Velocity (a.u./sec) |  |  |  |  |  |  |  |
| -Mean | 2.24x10<sup>-2</sup> | 2.51x10<sup>-2</sup> | 0.040 |  | NA | NA | NA |
| -CV (SD/mean) | 0.27 | 0.20 | 0.062 |  |  |  |  |
| **Dual task** | **iRBD**<br>**(n=32) \*** | **Controls**<br>**(n=27) \*** | **p-value**<br>**(unpaired)** | **p-value**<br>**(paired)** | **Sn** | **Sp** | **AUC**<br>**(95% CI); cut-off** |
| Amplitude (a.u.) |  |  |  |  |  |  |  |
| -Mean | 0.54 | 0.65 | 0.001 | <0.001<sup>a</sup>, 0.208<sup>b</sup> | 66.7% | 67.9% | 0.71 (0.59 to 0.85); 0.6 |
| -CV (SD/mean) | 0.32 | 0.22 | <0.001 | <0.001<sup>a</sup> 0.087<sup>b</sup> | 75.8% | 64.3% | 0.75 (0.63 to 0.87); 0.3 |
| Velocity (a.u./sec) |  |  |  |  |  |  |  |
| -Mean | 1.89x10<sup>-2</sup> | 2.23x10<sup>-2</sup> | 0.007 | 0.002<sup>a</sup>, 0.002<sup>b</sup> | 66.7% | 67.9% | 0.69 (0.55 to 0.83); 0.2 |
| -CV (SD/mean) | 0.38 | 0.28 | 0.023 | 0.002<sup>a</sup>, 0.003<sup>b</sup> | 66.7% | 67.9% | 0.67 (0.54 to 0.82); 0.3 |

Dual task: finger tapping + mental task. Amplitude was normalised. a.u.: arbitrary unit. Sn: sensitivity, Sp: specificity. \*Three finger tapping video recordings (one iRBD and two controls) were excluded due to technical issues related to hand image processing. P-value extracted from Welch's test for unequal variances; Unpaired t-test: iRBD vs controls, Paired t-test (whithin groups): a) iRBD single and dual task), b) paired t-test (controls single and dual task)

### Motor battery vs MDS-UPDRS-III

To minimize the risk of multicollinearity when combining motor markers in the multivariate logistic regression model required to undertake the ROC analysis, we investigated whether motor parameters were correlated one to each other. The AT-DFT was strongly correlated with the KS-DFT parameter (r=-0.77). Similarly, finger tapping amplitude and velocity were found to be strongly correlated (r=-0.77). The rest of motor markers (BRAIN test -KS, AT, IS-, walking time and handwriting tasks) could be considered independent (r<0.3). Thus, the multivariate logistic regression analysis did not include AT- DFT and amongst kinetic parameters of finger tapping we only selected CV amplitude.

The combination of the most salient motor markers (BRAIN (KS, AT, IS), DFT (KS, IS), % change in walking task, CV amplitude) was found to have 90.3% sensitivity for 89.3% specificity (AUC 0.94, 95% CI 0.88 to 0.99). The motor battery offered a higher accuracy than MDS-UPDRS-III, both the overall score (81.8% sensitivity for 72.4% specificity and AUC 0.83, 95% CI 0.72 to 0.93) and the MDS Task force research criteria (MDS-UPDRS-III score minus action tremor) (69.7% sensitivity for 72.4% specificity and AUC 0.81, 95% CI 0.71 to 0.91). The motor battery was also accurate to distinguish between iRBD with and without Subthreshold Parkinsonism (77.8% sensitivity for 86.4% specificity and AUC 0.91, 95% CI 0.80 to 1.00) (Table 4, Figure 3).

**Figure 3.**
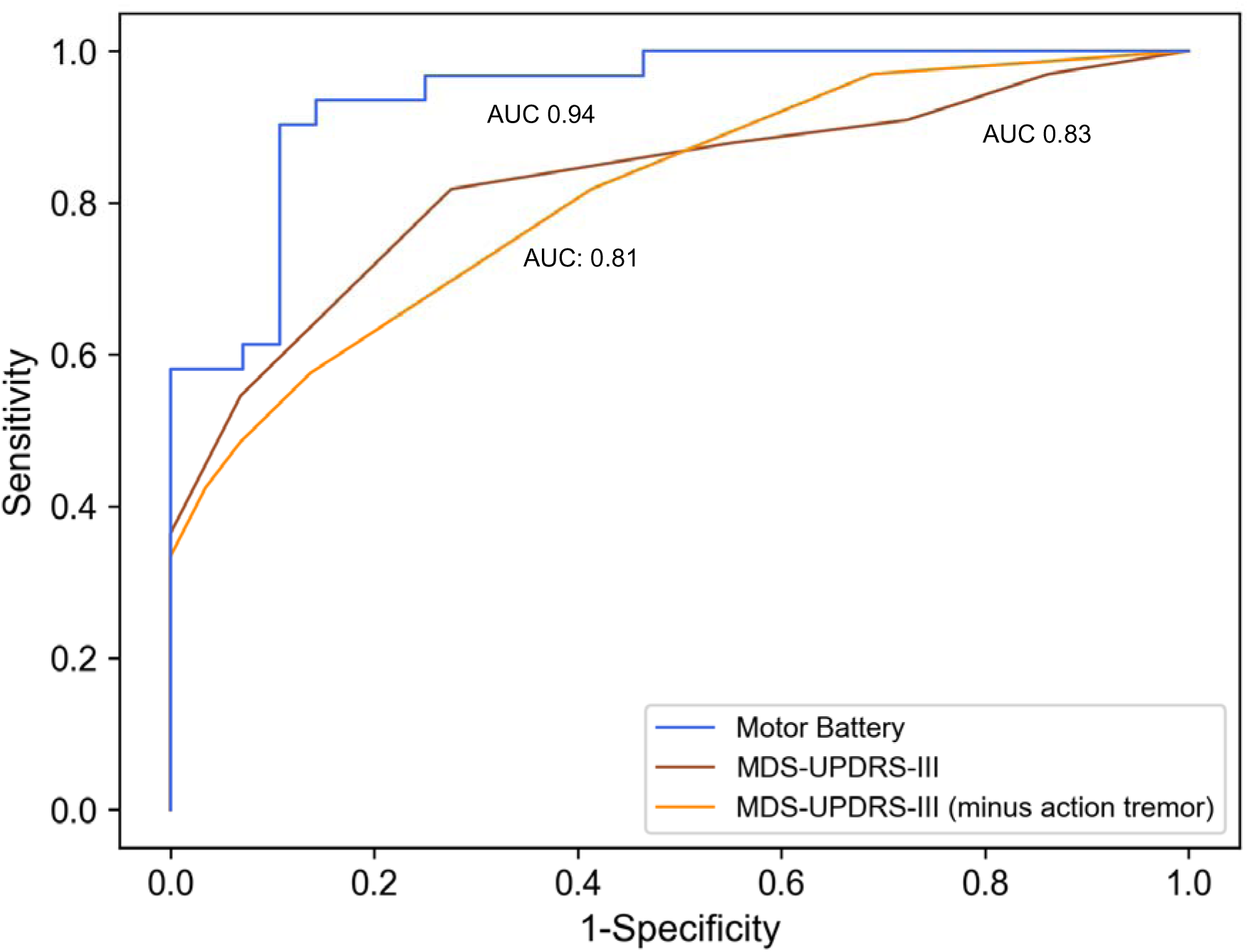
ROC curves for the motor battery (blue line, AUC 0.94), the total score of the MDS-UPDRS-III (brown line, AUC 0.83) and the MDS-UPDRS-III minus action tremor (MDS task force criteria for Subthreshold Parkinsonism) (orange line, AUC 0.81) to distinguish iRBD patients from controls

**Table 4.**
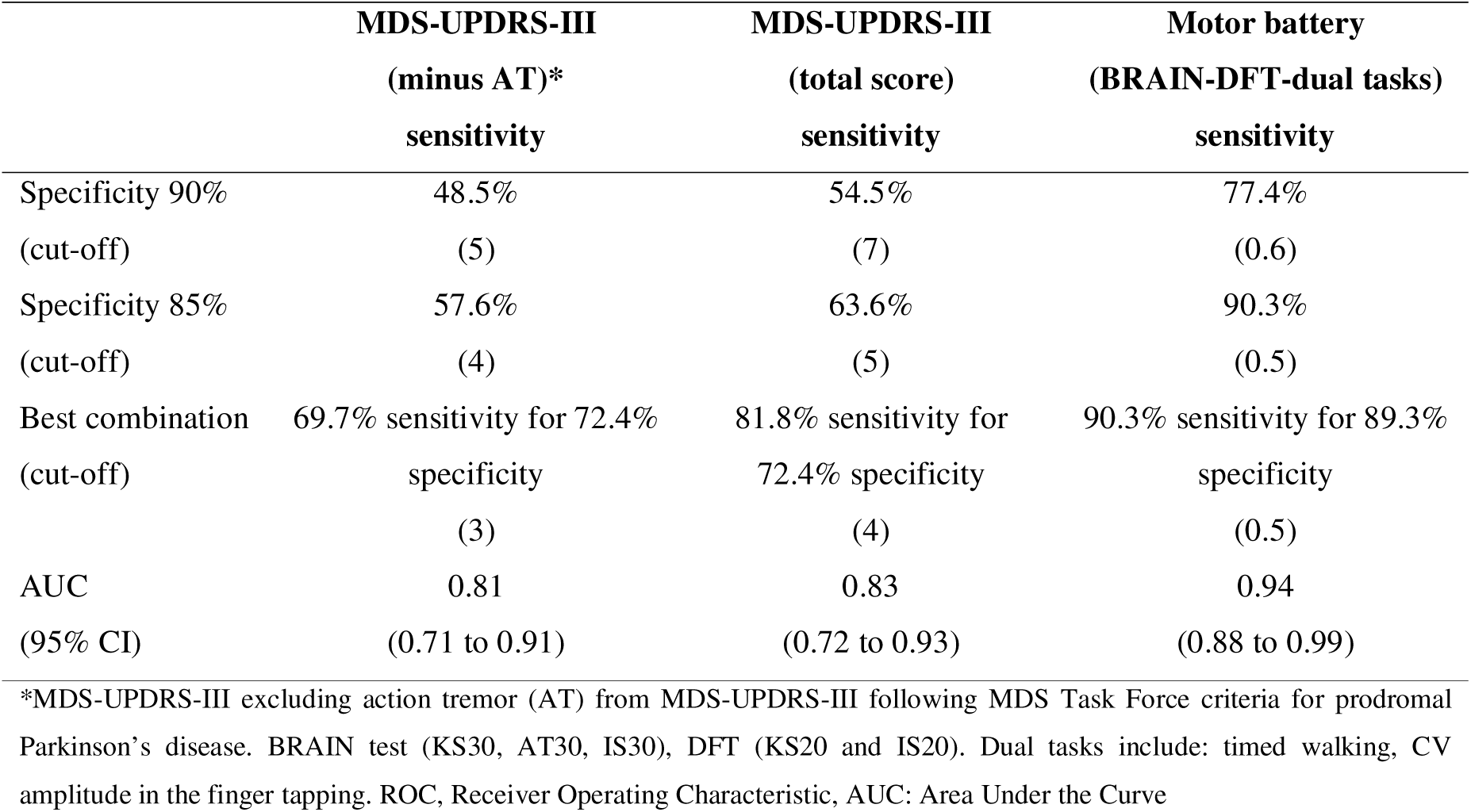
ROC analysis of motor battery vs clinical scales.

## Discussion

We have used a battery of motor tests which were tested together with the standardized MDS-UPDRS-III scale in people with v-PSG-confirmed iRBD and age- and sex-matched controls. Slow and erratic movement was the common denominator across all the motor tasks in the iRBD group. This is also the first study showing that dual tasking unmasked motor dysfunction in finger tapping. Both walking pace and finger tapping worsened in people with iRBD while performing a mental task, displaying a finger tapping with smaller amplitude and speed than controls.

There is evidence that some people with iRBD have motor dysfunction years before PD diagnosis.^19^ Higher motor scores in the MDS-UPDRS-III,^7, 20–23^ slow finger tapping,^7, 20, 21, 24, 25^ decreased gait velocity with greater step variability,^20, 26, 27^ and speech abnormalities^28^ are some of the anomalies described in the literature. Apart from supporting the existing evidence of mild parkinsonian signs being prevalent in iRBD, our study reveals subtle motor impairment in iRBD that had not been observed by the patients or noted on clinical examination. This is the case with the DFT and BRAIN test, as well as timed handwriting and dual tasking (mental task in combination with walking and finger tapping). Both tests distinguished iRBD from controls and were accurate in detecting individuals with Subthreshold Parkinsonism among the iRBD group. Having an online tool that can be administered remotely and accurate enough to detect people at risk with signs of subclinical parkinsonism will have important implications when neuroprotective treatments become available. This is particularly relevant based on a recent prospective multicenter study of 1160 subjects who were followed over an average of 3.3 ± 2.2 years.^19^ Authors used linear mixed-effect modelling to estimate annual rates of clinical marker progression. Motor features were found to progress faster and require the lowest sample sizes to detect differences in marker progression at 50% of drug efficacy.

Accounting for potential confounding factors such as age and sex is important to define the boundaries between neurodegeneration and natural variability driven by ageing process and sex-related differences. With that in mind, we compared the iRBD patients with controls matched for age and sex which provides stronger support that the motor anomalies seen in iRBD could be explained by a compensated neurodegenerative process. Lo and colleagues found that quantitative motor tools were more accurate and consistent in capturing motor change over time than standardized clinical scales.^20^ Moreover, Fereshtehnejad and collaborators concluded that slow alternate tap test had the longest period prior to diagnosis (8 years), followed by rigidity (3 years) and tremor (2 years).^29^ Arora and colleagues created a smartphone-based set of quantitative motor assessments including finger tapping, voice recording, balance and reaction time test and tremor analysis.^24^ Internal validation using machine learning showed that they were highly effective in discriminating between people with iRBD, PD patients and controls. In this study motor tasks were restricted to a single device used for data collection; therefore, it was not possible to extract detailed kinematic parameters of gait and finger tapping. Second, a high discrimination accuracy of a machine learning algorithm does not necessarily denote high clinical explanatory power given that sophisticated algorithms are mathematically complex, and therefore difficult to be interpreted from a clinical perspective. It is with that in mind that there is a need to develop a motor battery that can measure early motor dysfunction, not only in a quantitative manner, but also to be used in a replicable and interpretable way. To be replicable on a large scale, it is important to use inexpensive and user-friendly tools. To be interpretable, this battery needs to have a clinically meaningful explanation to avoid making spurious conclusions.

We found that a higher proportion of iRBD patients had Subthreshold Parkinsonism based on the MDS-UPDRS-III.^8, 30^ Findings that were supported by our motor battery of tests which raise the possibility that incoordination and slowing may be early signs of neurodegeneration. Rhythm disturbances have already been suggested as a early motor marker in iRBD.^31^ In line with finger tapping speed, we found that iRBD participants had slower handwriting, taking on average 10 seconds more to write three sentences than controls.

Finger tapping is another promising clinical biomarker of PD in people with iRBD. Several longitudinal studies have found that people with iRBD had slow finger tapping,^7, 20, 29^ reaching a sensitivity and specificity of 80% to identify the presence of iRBD who converted to PD or dementia.^2^ The Purdue pegboard test, a keyboard alternate tap,^32, 33^ and a 3D contactless motion capture of finger tapping are examples of methods used to capture early signs of upper limb bradykinesia. The alternate tap test was found to be one of the earliest motor signs in iRBD prior to PD diagnosis, showing 66.7% sensitivity for 77.3% specificity 2 years before clinical diagnosis, which decreased at a longer (6 years) prodromal interval down to 55% for the same degree of specificity. ^29^ In line with the study carried out by Fereshtehnejad and colleagues, we found that people with iRBD performed the BRAIN test less quickly than controls. The number of alternate taps per task in the BRAIN test alone had a slightly lower sensitivity than the alternate tap test used by Fereshtehnejad 2 years prior to phenoconversion (61% vs 66.7%). However, the online BRAIN test had a higher specificity (85% vs 77.3%) than the traditional alternate tap test. These findings support the notion that slow alternate finger tapping is an early motor marker with high prediction power of conversion to parkinsonism.

Finger tapping movements have also been assessed using the MDS-UPDRS-III instructions. Růžička and collaborators used a contactless 3D motion capture system to track the finger tapping task in the MDS-UPDRS-III.^25^ They tested 40 v-PSG-confirmed iRBD patients, 25 *de novo* PD patients and 25 healthy controls. They found that people with iRBD had a more pronounced decrement in the amplitude of finger tapping than controls. The instrumental analysis of finger tapping was able to distinguish iRBD from controls with 76% sensitivity and 63% specificity, which is comparable to the accuracy seen in the SMART test during dual tasking.

In a previous study, we tested a group of people with idiopathic anosmia using the SMART test.^18^ In common with the iRBD patients, in this study they performed the finger tapping task more slowly and with smaller amplitude than controls. Unlike patients with iRBD, however, these differences were seen under natural conditions. Most of the people with idiopathic anosmia in the trial had a normal finger tapping sub-score in the MDS-UPDRS-III, indicating that the SMART test could detect subtle motor signatures difficult to pick up with the naked eye. The DFT test mirrored the finger tapping task assessed in the MDS-UPSRS-III. Unlike the SMART test and 3D motion capture test, the DFT is a simple keyboard-based tap test. It can be used remotely, which facilitates its applicability on a large scale. In line with previous studies, people with iRBD had slower repetitive finger tapping rate than controls. In addition, higher incoordination appeared to be common in our iRBD group, not only when performing the DFT but also with the BRAIN test denoting a potential novel motor signature in iRBD.

In contrast with finger tapping, the effect of dual tasking on unmasking motor dysfunction has mainly been studied in gait.^34^ There is evidence suggesting that attention might have a role as a cognitive compensatory mechanism of motor dysfunction in posture control and gait.^35^ During the early stages of Parkinson’s disease it has been proposed that patients activate attention circuits to compensate for their motor dysfunction.^36, 37^ As disease progresses compensatory mechanisms fail with the onset of motor symptoms. Dual tasking might be able to disrupt these mechanisms by breaking the attention loops. The effect of dual tasking on gait has also been studied in healthy older people where no change occurred.^38^ In our study, both groups had similar age but differed in terms of cognitive function. Patients with higher cognitive burden might be more susceptible to challenging conditions, which could explain the differences seen between the effect of dual tasking on motor performance (walking speed and finger tapping) in iRBD patients compared with controls. A follow-up of these patients will have important prognostic implications, as it would be relevant to assess whether people who are more sensitive to dual tasking conditions are more prone to develop cognitive impairment in the future.

In the present study, observer bias in the MDS-UPDRS-III scoring could not be ruled out due to lack of a blinded assessment. To overcome this limitation, video recordings were examined by two movement disorder experts who were blinded to case/control status. This agreed with unblinded in-person assessment in 90.3% (56/62) participants. The clinical impression was also supported by other objective motor tools (e.g., keyboard-based tapping tests and handwriting speed). Another limitation derived from the cross-sectional nature of the study is the lack of information of motor changes over time. Follow up of this group will be crucial to know which markers if any predict a future diagnosis of PD or other neurodegenerative disorder. Previous literature found that the accuracy and predictive value of motor signs increased exponentially two years prior to PD diagnosis.^29, 39^ This suggests that motor dysfunction does not progress in a linear fashion and that motor change, might serve as a proximity marker of phenoconversion.^40^

To conclude, the tests used in this study were able to detect early patterns of motor dysfunction that are not included in standardized clinical scales. Speed, elements of erratic finger tapping movement and particular difficulty with dual tasking might be early motor anomalies in iRBD. Keyboard tapping was found to be slow and erratic in people with iRBD. Dual tasks unmasked hypokinetic finger tapping and slower walking pace.

## Data Availability

All data produced in the present study are available upon reasonable request to the authors

## Acknowledgment

all participants of the study

## Authors’ Roles

1. Research project: A. Conception, B. Organization, C. Execution
2. Statistical Analysis: A. Design, B. Execution, C. Review and Critique
3. Manuscript Preparation: A. Writing of the first draft, B. Review and Critique

CS: 1A, 1B, 1C. 2A, 2B, 2C. 3A, 3B

LPC: 1B, 2C, 3B

BFRH: 1B, 2C, 3B

HC: 2C, 3B

AG: 2C, 3B GL: 2C, 3B

AJL: 2C, 3B AS: 2C, 3B

AJN:1A, 1B. 2C. 3B

## Financial Disclosures (for the preceding 12 months)

GS: speaker fees for Takeda, consultancy fees for Eisai, and institutional income from Bioprojet and Jazz

AS reports having received research funding or support in the last 12 months from University College London, National Institute of Health (NIHR), National Institute for Health Research ULCH Biomedical Research Centre, the International Parkinson and Movement Disorder Society (IPMDS), the European Commission, Parkinson’s UK and the Economic and Social Research Council. Honoraria for consultancy from Abbvie; and license fee payments from the University College London. Royalties from Oxford University Press.

AJN reports grants from Parkinson’s UK, Barts Charity, Cure Parkinson’s, National Institute for Health and Care Research, Innovate UK, Virginia Keiley benefaction, Solvemed, the Medical College of Saint Bartholomew’s Hospital Trust, Alchemab, Aligning Science Across Parkinson’s Global Parkinson’s Genetics Program (ASAP-GP2) and the Michael J Fox Foundation. Prof Noyce reports consultancy and personal fees from AstraZeneca, AbbVie, Profile, Roche, Biogen, UCB, Bial, Charco Neurotech, uMedeor, Alchemab, Sosei Heptares and Britannia, outside the submitted work.

The rest of the co-authors do not have funding sources to report in the previous 12 months.

