## supplementary data for "The motor anomalies seen in isolated REM sleep behavior disorder"

**Supplementary material**

**Slow Motion Analysis of Repetitive Tapping (SMART) test**

Video analysis

We developed a software able to automatically detect the hand and process video images to analyse the finger tapping task. Due to the variability in the video acquisition, hand images were resized, rotated, and flipped to have a homogeneous sample. The videos were processed and analysed together with the collaboration of a bioinformatic with experience using Python programming language and OpenCV^1^ library. A 2D Convolutional Neural Network (CNN) was trained to detect eight key landmarks of the first and the second fingers (Figure 4.2). A total of 3934 randomly extracted frames among all the videos were manually labelled and were used as a dataset for the CNN training. The complete dataset was divided into a train and a test dataset using a ratio of 0.8:0.2 respectively. The CNN was implemented in PyTorch.^2^ Videos were then processed, and eight key hand landmarks were detected in every frame along the video using the trained CNN. In order to study the fine movement of the tapping task, the distance between the distal part of the first and the second fingers was computed through the video (first and last key landmarks). The computed distances are not real-distances, and they were normalised to be comparable between samples. This step limits the power of the technique since the absolute opening of the hand cannot be seen due to the normalisation process. To overcome this limitation, the angle formed between the distal part of the first and second finger and the key landmark corresponding to the metacarpal joint was also computed (Figure 4.2). This value can give information about the absolute opening of the hand during the tapping task and gives a useful variable to be compared among participants.

Maximum amplitude peaks were detected at each tap and linear regression models were fitted to those signal peaks. Frequencies were measured as number of taps per second. Velocities were calculated as the first derivative of the signal, and a similar process was applied to obtain the peaks of maximum velocities over time. The integrals of the signals were also computed. This value gives a measure of the freezing of the hand during the tapping task. All the signal processing was done using SciPy^3^ and NumPy^4^ libraries.

I analysed three kinetic parameters

1. Amplitude: angle formed between index finger and thumb
2. Frequency: number of taps per second
3. Velocity: distance travelled per second extracted from the derivative of the amplitude.

For each parameter, we calculated the mean, SD and Coefficient Variation (CV) (SD/mean).

**Figure** SMART test hand detection: 8 key landmarks across the first and the second finger (red). Angle between 1-4-8 key landmarks (black). Extrapolated amplitude between point 1 and 8 (blue)

a. Dominant hand of controls with the most affected side in the PD group
b. Non-dominant hand of controls with the most affected side in the PD group
c. Dominant hand of anosmia with dominant hand of controls
d. Dominant hand of anosmia with asymptomatic side of patients with unilateral PD.

ROC curves were drawn to find the optimal cut-off value which maximises the combination of sensitivity and specificity (Youden’s J index) for SMART test parameters separately and in combination. I used Spearman’s correlation coefficient to correlate SMART test parameters (continuous) with FT-sub-scores from the MDS-UPDRS-III (ordinal). Since I ran multiple hypothesis tests – one for each component of the test parameters (mean, CV, and slope) – I selected a more stringent cut-off for the level of significance.
